# Phase II Study of Myeloablative 8/8- or 7/8-Matched Allotransplantation with Post-Transplant Cyclophosphamide, Tacrolimus, and Mycophenolate Mofetil: Marked Reduction in GVHD Risk Without Increased Relapse Risk Compared to Historical Cyclosporine/Methotrexate

**DOI:** 10.1101/2023.03.24.23287521

**Authors:** Najla El Jurdi, Alex Hoover, Daniel O’Leary, Qing Cao, Ashish Gupta, Christen Ebens, Joseph Maakaron, Brian C. Betts, Armin Rashidi, Mark Juckett, Troy Lund, Veronika Bachanova, Margaret MacMillan, Jeffrey Miller, Paul Orchard, John Wagner, Gregory Vercellotti, Daniel Weisdorf, Kathryn Dusenbery, Stephanie Terezakis, Shernan Holtan

**Author notes:** Authors Contributed Equally (Najla El Jurdi, Alex Hoover, Daniel O’Leary).

## Abstract

**Introduction:** Graft-versus host disease (GVHD) is a major limitation to the success of allogeneic hematopoietic cell transplant (HCT). We hypothesized that the GVHD prophylaxis regimen of post-transplant cyclophosphamide (PTCy), tacrolimus (Tac) and mycophenolate mofetil (MMF) would reduce the incidence of GVHD in patients receiving a matched or single antigen mismatched HCT without an increase in risk of malignant relapse.

**Methods:** This is a phase II study conducted at the University of Minnesota using a myeloablative regimen of either: (A) total body irradiation (TBI, total dose 1320 cGy, administered in 165 cGy fractions, twice a day from days -4 to -1) or (B) Busulfan 3.2mg/kg daily (cumulative AUC 19,000 – 21,000 μmol/min/L) plus fludarabine 160mg/m^2^ days -5 to -2, followed by a GVHD prophylaxis regimen of PTCy (50mg/kg days +3 and +4), Tac and MMF (beginning day +5). The primary endpoint is cumulative incidence of chronic GVHD requiring systemic immunosuppression at 1-year post-transplant. We compared results to our previous myeloablative protocol for matched donors utilizing cyclosporine/methotrexate (CSA/MTX) GVHD prophylaxis.

**Results:** From March 2018 - June 2022, we enrolled and treated 125 pediatric and adult patients with a median follow up of 472 days. Grade II-IV acute GVHD occurred in 16% (95% confidence interval (CI): 9-23%); Grade III-IV acute GVHD was 4% (CI: 0-8%). No patients experienced grade IV GVHD, and there were no deaths due to GVHD before day 100. Only 3 developed chronic GVHD requiring immune suppression, (4%, CI: 0-8%). Two-year overall survival (OS) was 80% (CI: 69-87%), and (graft-versus-host disease-free, relapse-free survival) GRFS 57% (CI: 45-67%), both higher than historical CSA/MTX. The incidence of grade II-IV aGVHD, cGVHD, and NRM were all lower with PTCy/Tac/MMF compared to historical CSA/MTX. One-quarter (25%) experienced relapse (CI: 15-36%) similar to historical CSA/MTX. There was no statistically significant difference in survival outcomes between recipients of matched versus 7/8 donors.

**Conclusion:** Myeloablative HCT with PTCy/Tac/MMF results in extremely low incidence of severe acute or chronic GVHD, the primary endpoint of this clinical trial. Relapse risk is not increased compared to our historical CSA/MTX cohort.

## Introduction

Graft-versus-host disease (GVHD) has been the major complication limiting success of allogeneic hematopoietic cell transplantation since the field began 70 years ago. The standard regimen to prevent GVHD, a combination of calcineurin inhibitor and methotrexate, has been used since the 1980s, but it has significant tissue toxicity and does not decrease the risk of chronic GVHD (reviewed in ^1^). The drug abatacept was approved by the FDA for the prevention of acute GVHD in 2021, however it also does not address the risk of chronic GVHD. ^2^ Intensifying broad immunosuppression using ATG or other agents may decrease the occurrence of GVHD but simultaneously increase the risk of relapse or infections with no overall net benefit.^3, 4^ Post-transplant cyclophosphamide-based regimens are increasingly being tested for GVHD control, with an evolved understanding of its mechanism of action including restraint of alloreactive conventional T cell proliferation function with preferential recovery of regulatory T cells.^5, 6^ In a single center, prospective phase II study, we examined the hypothesis that using the regimen of PTCy, tacrolimus (Tac), and mycophenolate mofetil (MMF) would lower the incidence of both acute and chronic GVHD without increasing the risk of relapse. We compared the results of this regimen to a historical group of allogeneic hematopoietic cell transplant recipients who received well-matched allografts and myeloablative conditioning (MAC) with cyclosporine and methotrexate (CSA/MTX) prophylaxis at our institution to understand the differences in GVHD, relapse, and survival outcomes with PTCy/Tac/MMF compared to our historical standard.

## Methods

This is an analysis of a phase II study at the University of Minnesota using a MAC regimen of either: 1. total body irradiation (TBI, total dose 1320 cGy administered twice a day from days -4 to -1) or 2. Busulfan 3.2mg/kg daily (cumulative AUC 19,000 – 21,000 μmol/min/L) plus fludarabine 160mg/m^2^ days - 5 to -2 for patients unable to receive radiation, followed by a GVHD prophylaxis regimen of PTCy (50mg/kg days +3 and +4), Tac and MMF (both beginning day +5). The primary endpoint was cumulative incidence of chronic GVHD requiring systemic immunosuppressive treatment (IST) at 1-year post-transplant. This clinical trial is registered at ClinicalTrials.gov under NCT03314974. Eligibility included age ≤ 60 years, hematologic malignancy with candidacy for allotransplantation, matched related donor (MRD) or unrelated (URD) donor with either a bone marrow (BM) or filgrastim-mobilized peripheral blood (PB) graft. This trial was a prospective clinical trial reviewed and approved by the Masonic Cancer Center Protocol Review Committee and Human Subjects Institutional Review Board at the University of Minnesota. All patients signed Institutional Review Board–approved informed consent in accordance with the Declaration of Helsinki.

We compared the results from the current study to our previous MAC protocol which utilized a regimen of either cyclophosphamide (60mg/kg days -6 and -5)/TBI (1320 cGy administered twice a day from days -4 to -1) or busulfan (0.8 mg/kg/dose IV in 4 daily doses days -9 to -6 [total 12.8 mg/kg])/ cyclophosphamide (50 mg/kg days -5 to -2), with GVHD prophylaxis utilizing cyclosporine and methotrexate. The historical comparison cohort was also a prospective phase II clinical trial registered under NCT00176930. Patients underwent allogeneic HCT on this clinical trial between 2005 – 2018. Eligibility criteria regarding organ function and hematologic malignancy status were the same for each study, although the upper age limit was lower (age 55) for the historical cohort. Disease risk was classified as standard or high risk based on the American Society for Blood and Marrow Transplantation Request for Information 2006 risk scoring schema (http://www.asbmt.org).

For analysis of key clinical endpoints, the cumulative incidence estimator was used to calculate the probabilities of acute GVHD, chronic GVHD, relapse or disease progression, and engraftment, reflecting the non-event deaths as a competing risk.^7^ Similarly, cumulative incidence was used to estimate transplant related mortality (TRM) treating relapse as a competing risk. Fine and Gray regression analyses was used to compare the differences between cumulative incidence curves.^8^ The cut off significance level was 0.05, and all reported P values are two-sided. SAS 9.4 (SAS Institute, Inc., Cary, NC) and R version 4.1.2 (The R Foundation for Statistical computing) were used for all analyses.

## Results

From 2018 - 2022, we treated 125 patients with MAC and PTCy/Tac/MMF prophylaxis, with a median follow up of 472 days post-transplant. Demographics and baseline characteristics of this prospective cohort as well as the historical comparison cohort of patients receiving myeloablative transplantation with CSA/MTX are detailed in Table 1.

**Table 1.**
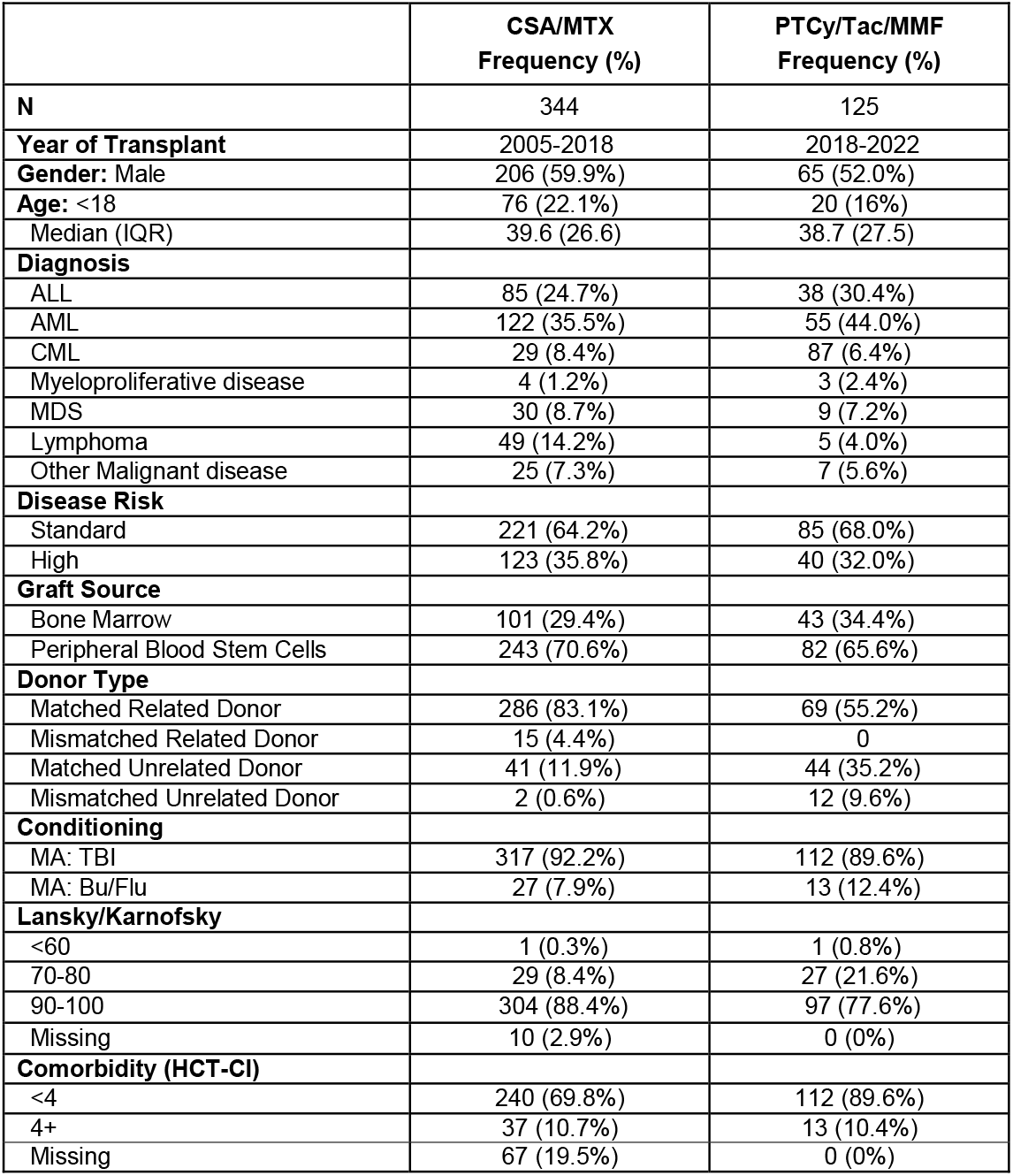
Patient baseline characteristics.

The historical CSA/MTX cohort was similar to the prospective PTCy cohort. Both groups had a slight male predominance (59.9% vs 52% for PTCy), similar proportion of pediatric patients (22.1% vs 16% for PTCy), and similar median age of 39.6 years versus 38.7 years for PTCy. The primary diseases for which transplant was performed were similar except for a slightly higher proportion of patients with lymphoma undergoing HCT with CSA/MTX (14.2% versus 4.0% with PTCy). Disease risk was similar between cohorts (high risk in 35.8% of patients versus 32.0% with PTCy). Graft sources were similar between the cohorts, except the CSA/MTX cohort utilized some mismatched related donors as the graft source (4.4% versus 0% for PTCy), and the PTCy cohort utilized more mismatched unrelated donors (9.6% versus 0.6% for CSA/MTX). Most recipients were conditioned with TBI (92.2% versus 89.6% for PTCy). There was a slightly higher proportion of patients with a performance status 90-100 (88.4% versus 77.6% with PTCy). The median time to neutrophil engraftment was 16 days for both cohorts, achieved in 97% receiving CSA/MTX (range 3-31 days) and 99% receiving PTCy (range 13-30 days). The median time to platelet engraftment was also the same at 26 days in both cohorts, achieved in 85% receiving CSA/MTX (range 0-372 days) and 95% in patients receiving PTCy (range 14-98 days). The median duration of hospitalization was 3 days longer in patients receiving PTCy, 28 days (interquartile range [IQR] 9 days) versus 25 days in patients receiving CSA/MTX (IQR 12 days).

GVHD endpoints are plotted with PTCy in comparison to the historical CSA/MTX population in Figure 1. Regarding the prospective study’s primary endpoint, only 3 of 105 patients developed chronic GVHD requiring systemic immune suppression at 1 year, for a cumulative incidence of 4% (CI: 0-8%). The cumulative incidence of Grade II-IV acute GVHD by day +100 was 16% (95% confidence interval (CI): 9-23%); Grade III-IV acute GVHD was 4% (CI: 0-8%). Zero patients experienced grade IV GVHD within 100 days after PTCy. All GVHD outcomes were statistically significantly worse with historical CSA/MTX (Figure 1), including grade II-IV aGVHD at 37% (CI 41-42%), grade III-IV aGVHD at 15% (CI 11-19%), and chronic GVHD requiring immunosuppression at 34% (CI 28-39%).

**Figure 1.**
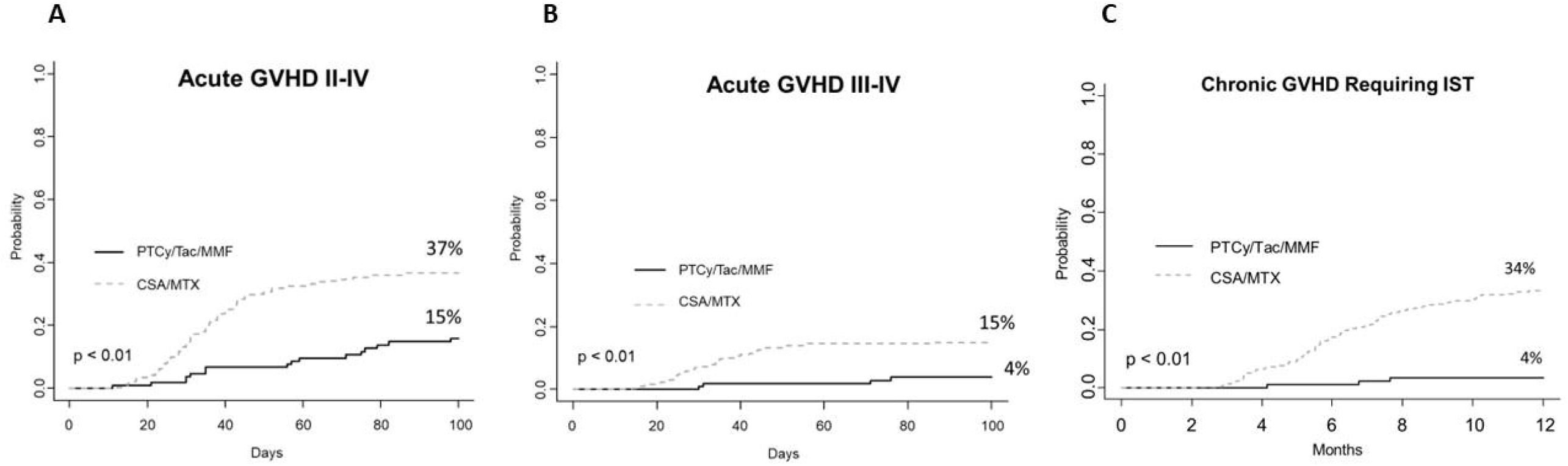
GVHD-related outcomes of PTCy/Tac/MMF versus historical CSA/MTX: (A) Acute GVHD grade II-IV, (B) Acute GVHD grade III-IV, and (C) Chronic GVHD requiring systemic immunosuppression.

Non-relapse mortality at 1 year (Figure 2A) was higher with CSA/MTX at 19% (CI 14-23%) than PTCy/Tac/MMF (10%, CI 4-16%, p=0.03). The two-year cumulative incidence of relapse was 25% (CI: 15-36%), which did not differ from the historical cohort at 21% (CI 16-26%, Figure 2B). However, with PTCy, we noticed a higher relapse probability in children ages 18 and younger (49%, 95% CI 23-74%) compared to children receiving CSA/MTX (26%, 95% CI 16-37%, p=0.03). There was no statistically significant difference in relapse rates with PTCy/Tac/MMF vs CSA/MTX in those over age 18 (p=0.99). Two-year OS was better with PTCy/Tac/MMF at 80% (CI: 69-87%) versus 64% (CI 58-69%, p<0.01, Figure 2C). Two-year GRFS was also better with PTCy/Tac/MMF at 57% (CI: 45-67%) than 25% (CI 20-30%, p<0.01, Figure 2D) with historical CSA/MTX.

**Figure 2.**
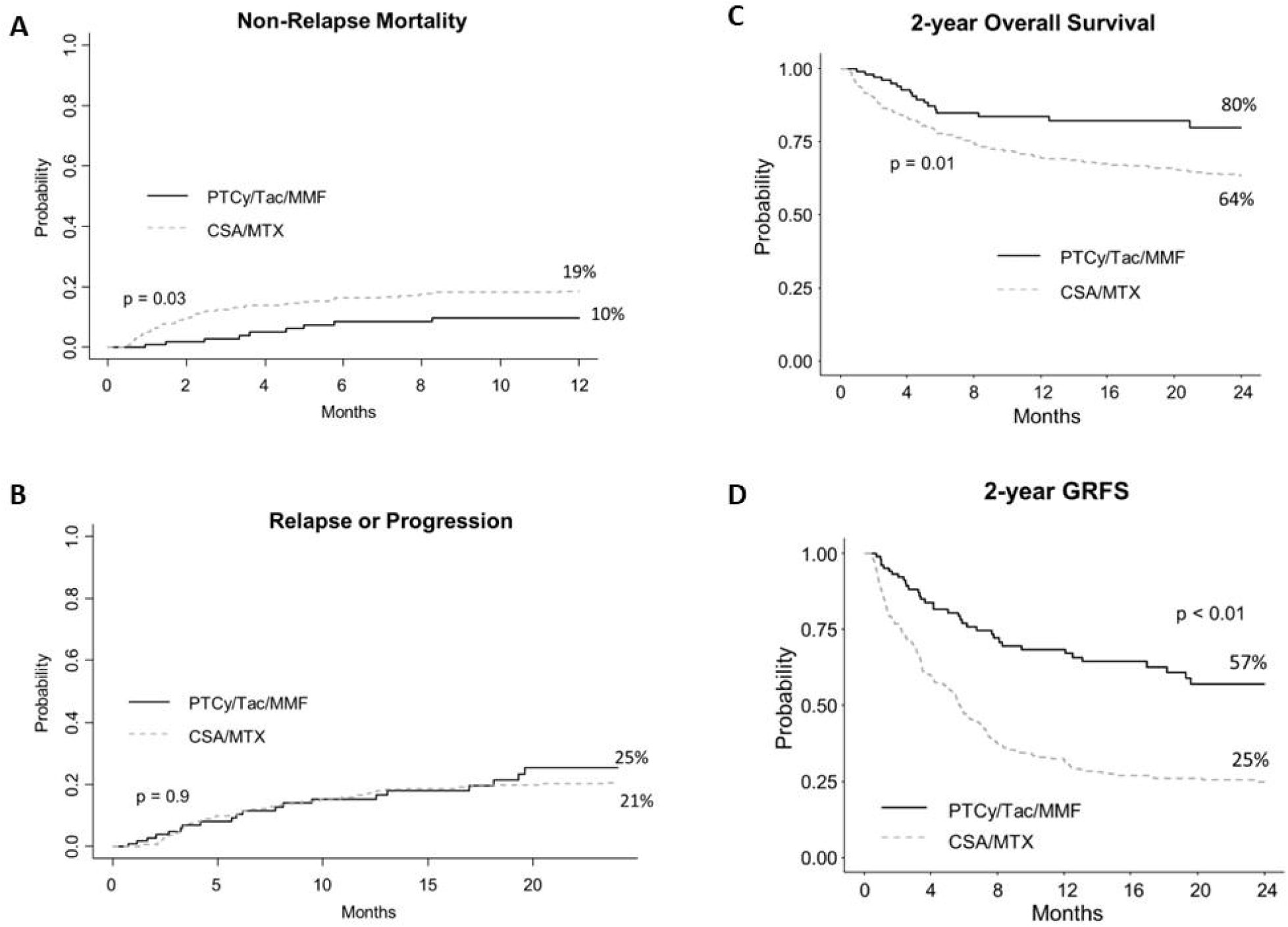
Relapse and survival outcomes of PTCy/Tac/MMF versus historical CSA/MTX: (A) Non-relapse mortality, (B) relapse/progression, (C) overall survival, and (D) graft-versus-host disease-free, relapse-free survival (GRFS).

In a multivariate analysis of 2-year OS of the cohorts (Table 2), PTCy/Tac/MMF was associated with an over 50% reduction in the hazard of death. High risk disease was also independently associated with a 52% higher hazard of death compared to those with standard risk disease. Patients over age 40 had an 87% higher hazard of death compared to patients under the age of 18.

**Table 2.** Multivariate analysis of 2-year OS.

| Parameter | Group | Hazard Ratio CI 95% | P-value |
| --- | --- | --- | --- |
| GVHD Prophylaxis | CSA/MTX | 1.00 | <0.01 |
|  | PTCy/Tac/MMF | 0.49 (0.29-0.81) |  |
| Age (y) | <18 | 1.00 | 0.06 |
|  | 18-39 | 1.49 (0.85-2.63) |  |
| | $\geq 40$ | 1.87 (1.11-3.13) | |
| Disease Risk | Standard | 1.00 | 0.02 |
|  | High | 1.52 (1.06-2.18) |  |

Within the PTCy/Tac/MMF cohort, we performed exploratory analyses and found no difference in overall survival between HLA 7/8 vs 8/8 matches (p=0.99), sibling versus unrelated donor graft (p=0.33), the use of marrow versus peripheral blood stem cells (p=0.79), disease status at transplant, including the presence or absence of measurable residual disease (4 patients with MRD in this cohort, p=0.16), or type of conditioning (Bu/Flu vs TBI, p=0.21). Causes of death varied by donor source (Supplemental Figure 1). Relapse accounted for the most deaths in sibling transplants (71% of sibling HCT deaths) and the second leading cause of death for unrelated donor transplants (33% of URD HCT deaths). The leading cause of death in unrelated donor transplants was respiratory failure due acute respiratory distress syndrome or idiopathic pneumonia syndrome (44% of URD HCT deaths). Only one death attributed to GVHD occurred in the PTCy cohort in a patient with fatal late-onset stage 4 liver acute GVHD that occurred after tacrolimus taper.

## Discussion

The results of this phase II study show that GVHD prophylaxis with PTCy/Tac/MMF with a well-matched donor and a MAC regimen based on TBI or Busulfan/Fludarabine results in a notably low (4%) incidence of chronic GVHD requiring IST at one year, which was the primary endpoint of the trial. It is noteworthy that this regimen is both feasible and effective for both pediatric and adult patients up to the age of 60.

The survival advantage derived from this protocol compared to historical CSA/MTX is attributed to the marked decrease in both severe acute and chronic GVHD without any apparent increase in the risk of relapse in adult patients. However, it must be acknowledged that there was a higher rate of relapse observed in pediatric patients receiving PTCy/Tac/MMF when compared to CSA/MTX, thus a careful consideration of the balance between GVHD risk and relapse risk is advisable in pediatric HCT. While we acknowledge several advancements in supportive care may have contributed to improved survival in patients transplanted in 2018-2022 versus 2005-2018, the differences in GVHD-specific outcomes are striking and likely the primary driver of the results.

PTCy was first developed clinically in the haploidentical setting.^9, 10^ Subsequent results have been encouraging in studies of PTCy-based GVHD prophylaxis in the context of well-matched donor MAC allotransplantation. A 3-site prospective study of single-agent PTCy in matched bone marrow transplantation and MAC demonstrated a relatively low grade of grade III-IV aGVHD at 15% and chronic GVHD at 14%, with long-term follow up showing a low global IST burden in recipients of PTCy.^11, 12^ A subsequent study to expand these results to the adult peripheral blood stem cell MAC transplant setting found that addition of CSA to PTCy maintained low rates of chronic GVHD requiring IST at 1 year at 16%.^13^ Children and young adults also have a notably low rate of chronic GVHD after PTCy, with reported incidence rates as low as 0% after matched transplantation and 5-14% with haploidentical grafts.^14-16^ Our observed rates of chronic GVHD requiring IST at 1 year are low at 4% in the overall cohort, which may be owed to a combination of factors, including the use of Tac over CSA, the addition of MMF, and the allowance of bone marrow as a graft source in this protocol. Given the relative ease of procurement of PBSC versus bone marrow, the use of PTCy may markedly reduce the need for bone marrow harvesting in the future.

The regimen of PTCy/Tac/MMF used in this study has recently been proposed as the new standard of care for well-matched reduced intensity peripheral blood stem cell transplants based upon the results of phase III BMT CTN 1703. (Bolaños-Meade et al, NEJM, manuscript accepted). Our observed rates of grade III-IV acute and chronic GVHD requiring IST at 1 year are lower than that of 1703 (6.3% and 12.5% respectively), also likely due to the allowance of children and bone marrow grafts on this protocol. A randomized phase III study similar to BMT CTN 1703 has not yet been completed in the myeloablative setting; BMT CTN 1301 utilized MAC but BM as the graft source and single agent Cy, thus results are not directly comparable.^17^ Co-stimulation blockade with abatacept has been studied in the MAC setting, but rates of chronic GVHD remain high at >50%.^2^ Thus, a PTCy-based regimen may be preferable to Tac/methotrexate/abatacept in view of lower chronic GVHD risk, although this has not been directly tested in a randomized study.

Prevention of relapse/progression of the underlying disease should likely be the next step in the evolution of PTCy-based platforms. Novel enhancement of natural killer cell function or prevention of T cell exhaustion appears may be promising in the PTCy context.^18^ The approach currently under study at the University of Minnesota is addition of an oral aurora kinase A inhibitor to PTCy/sirolimus to preserve generation of antitumor cytotoxic T lymphocytes^19^; GVHD and relapse prevention are dual primary endpoints of this ongoing study (NCT05120570). Shared targets of GVHD and malignancy exist, such as CD83, which could be exploited to control both alloreactivity and enhance GVL in future studies.^20^

In summary, a GVHD prophylaxis regimen of PTCy/Tac/MMF shows significantly better acute and chronic GVHD control, without an increase in relapse risk, compared to historical CSA/MTX in well-matched MAC transplantation. The benefit of GVHD control is highest in adults. Pediatric HCT recipients may require a risk adapted approach to mitigate higher rates of relapse.

## Data Availability

Requests for data may be emailed to.

## Acknowledgements

This research was supported by NIH grant P30 CA77598 utilizing the Biostatistics Core of the Masonic Cancer Center, University of Minnesota and the National Center for Advancing Translational Sciences of the National Institutes of Health Award Number UL1TR002494. We would like to acknowledge our excellent team of BMT pharmacists, without whom this study would not be possible.

## Figure Legends

**Supplemental Figure 1: Causes of death by donor with PTCy/Tac/MMF GVHD prophylaxis**.

