## Supplementary figures and images for "Phase II Study of Myeloablative 8/8- or 7/8-Matched Allotransplantation with Post-Transplant Cyclophosphamide, Tacrolimus, and Mycophenolate Mofetil: Marked Reduction in GVHD Risk Without Increased Relapse Risk Compared to Historical Cyclosporine/Methotrexate"

### Figure S1

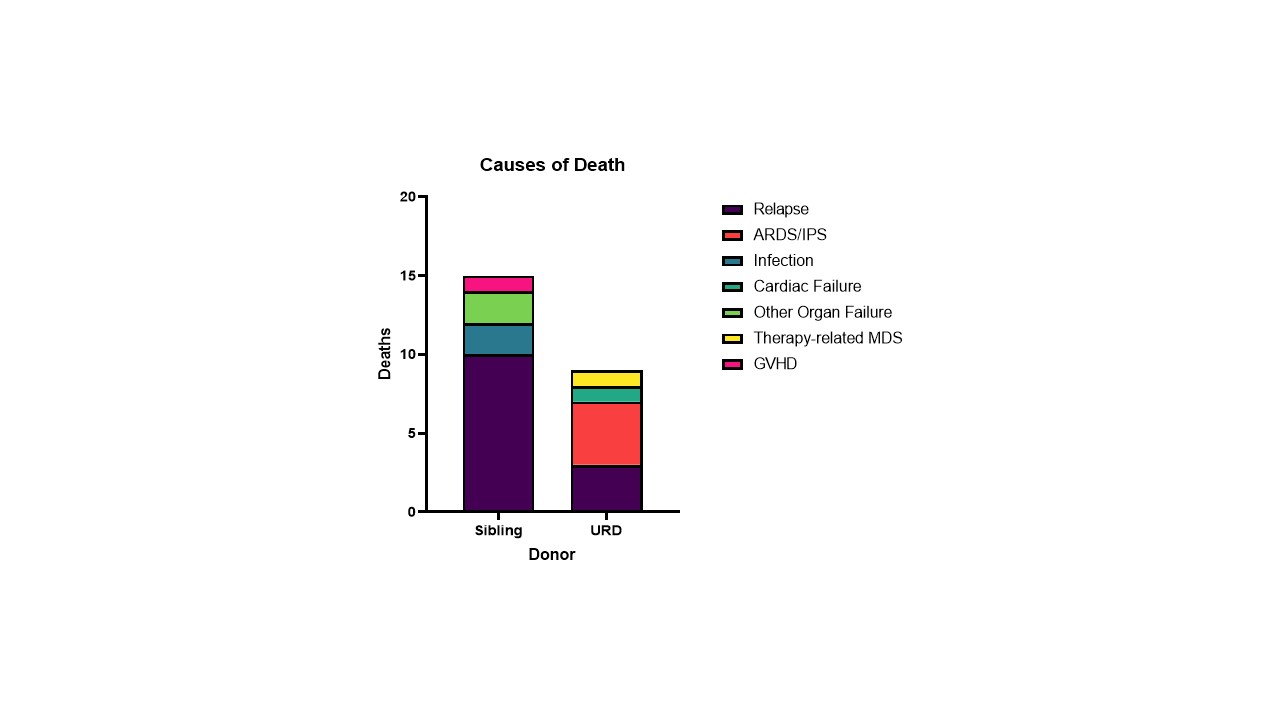
